# Epstein-Barr virus and its prognostic value in a cohort of Peruvian women with cervical cancer

**DOI:** 10.1101/2020.08.04.20167841

**Authors:** Denisse Castro, Juana Vera, Percy Soto-Becerra, Marco López-Ilasaca, Alejandro Yabar, Anais Cámara, Ana Fernández, Luis Malpica, Brady Beltrán

**Affiliations:** Department of Oncology and Radiotherapy, Hospital Nacional Edgardo Rebagliati Martins, Lima, Peru; Centro de Investigación de Medicina de Precisión, Universidad de San Martin de Porres, Lima, Peru; Department of Pathology, Hospital Nacional Edgardo Rebagliati Martins, Lima, Peru; Instituto de Evaluación de Tecnologías en Salud e Investigación – IETSI, EsSalud. Lima, Peru; Department of Medicine, Brigham and Women’s Hospital, Harvard Medical School, Boston, MA, USA; Department of Lymphoma and Myeloma, The University of Texas MD Anderson Cancer Center, Houston, TX, USA

**Keywords:** Epstein-Barr virus_1_, prognostic factor_2_, cervical cancer_3_, Peruvian women_4_, overall survival_5_

## Abstract

**Aim:** We aim to evaluate the prognostic effect of Epstein-Barr virus (EBV) infection on overall survival (OS) in Peruvian women with cervical cancer.

**Methods:** We conducted a retrospective cohort study. Polymerase chain reaction technique was used in paraffin-embedded tumor tissue for the detection of EBNA-1 and LMP-1. We used a multiple Cox proportional-hazard regression to estimate adjusted hazard ratios (aHR) for death and 95% confidence intervals (95% CI). In order to model continuous variables without categorization, we used a multivariable fractional polynomial approach. We performed a stability analysis using bootstrapping for internal validation.

**Results:** A total of 99 patients with cervical cancer were included. The prevalence of EBV in cervical cancer specimens was 22.2% (n=22). The 1-year and 5-year OS rates were 81.8% (95% CI 58.5-92.8) and 45% (95% CI 23.9-64.1) in the EBV-positive group compared to 78.8% (95% CI 67.7-86.4) and 37.8% (95% CI 25.7-49.8) in the EBV-negative group, respectively. In the multivariate analysis, positive EBV status was an independent prognostic factor for improved OS (aHR: 0.32; 95% CI 0.16 to 0.67; p=0.002) compared to negative EBV status.

**Conclusions:** EBV status is an independent prognostic factor for OS in cervical cancer. Evaluation of EBV status could be used as a clinical prognostic biomarker and to improve currently available prognostic models such as the FIGO system. Future prospective studies will be needed to validate these theories.

## INTRODUCTION

Cervical cancer (CC) is the third cause of cancer in the female population and the fourth leading cause of death worldwide (1,2). In Peru, CC is the second cause of cancer corresponding to 23.2% of the general population, and the third cause of death among Peruvian women (1).

In 2018, it was estimated that infections were responsible for 15% of cancers worldwide with 2.2 million (13%) of all cases associated to well-described infections (3). Human papillomavirus (HPV) infection is an indispensable requirement for the genesis of CC, however, less than 1% will develop cancer indicating that HPV is necessary but not sufficient for the development of carcinogenesis (4). In Peru, HPV strains 16 and 18 are responsible of most high-grade lesions and CC cases (5). Several biological and environmental factors have been implicated in the development of CC, some of them responsible for persistent HPV infection. Among those factors, Epstein-Barr virus (EBV) infection has been one of the most widely-studied viruses associated to the carcinogenesis of CC (6–9).

EBV was the first virus to be recognized as an oncovirus and has been linked to the development of various lymphoproliferative disorders (10), and solid tumors such as gastric carcinoma and nasopharyngeal carcinoma (NPC) (11,12). The association between EBV infection and CC was first reported in 1995 and since then several advances have been made (6). EBV DNA has been frequently found in vaginal and urethral fluids, suggesting the possibility of sexual transmission (7,13,14). Previous studies have reported higher EBV viral protein expression in invasive carcinomas compared to intraepithelial lesions or normal tissue (7,15). EBV expression is more commonly found in squamous histologies and in the presence of EBV/HPV co-infection (7,13,16). Different mechanisms have been postulated in regards to the synergic effect HPV and EBV co-infection exerts on tumor development and progression (17). Furthermore, the impact of EBV infection on survival in cancer patients is well documented with poor prognosis noticed in patients with lymphoma (10,18), as opposed to patients with certain carcinomas such as gastric and nasopharyngeal carcinomas (11,12).

Currently, there is a lack of real-world data regarding the role that EBV infection in the development of CC and its impact on survival. Brazil is the only Latin American country that reported a small number of cases on EBV and CC, however, no survival data has been yet presented (8). In Peru, there are no studies addressing the relationship between EBV infection and CC. Therefore, this study aims to evaluate the impact of EBV infection on survival in Peruvian women with cervical cancer.

## MATERIAL AND METHODS

### Study design and data source

A historical cohort of patients with anatomopathological diagnosis of squamous cell carcinoma of the cervix was identified and reviewed retrospectively through a search of clinical records at the Department of Oncology and Radiotherapy at the Edgardo Rebagliati Hospital in Peru. Patients were diagnosed between December 2013 to June 2014 and followed up until July 2019. The anatomopathological diagnosis of CC followed the histopathological criteria defined in the World Health Organization Classification (19). Additional inclusion criteria are: (1) patients must be ≥ 18-years-old; (2) anatomopathological diagnosis of CC should have been done (or reviewed and confirmed by our pathologist if biopsy was outside of our hospital) by the Department of Pathology at the Edgardo Rebagliati Hospital; and (3) only stages IIB, IIIA, and IIIB cervical cancer were considered for inclusion. We excluded patients with prior treatment in other healthcare centers; lost or destroyed medical records; incomplete or insufficient data for pathological characterization; and patients with insufficient material in cervical tissue to perform the qualitative real-time PCR testing for EBV analysis. We obtained the approval by the Institutional Review Board and Ethic Committee of the Hospital Nacional Edgardo Rebagliati Martins-EsSalud for the use of CC samples and data. This data was securely and anonymously stored. Supplementary Table 1 displays the REMARK profile describing key aspects of this study design and analysis.

**Table 1.** Characteristics of the total study cohort and according to EBV.

| Variables | All patients<br>(n = 99) | EBV-negative<br>(n = 77) | EBV-positive<br>(n = 22) | P value* |
| --- | --- | --- | --- | --- |
| Age, years† | 55.1 ± 15.7 | 55.4 ± 15.5 | 53.8 ± 16.4 | 0.675§ |
| Age group, n (%) |  |  |  |  |
| 28-34 | 8 (8.1) | 6 (7.8) | 2 (9.1) | 0.602 |
| 35-45 | 24 (24.2) | 17 (22.1) | 7 (31.8) |  |
| 46-92 | 67 (67.7) | 54 (70.1) | 13 (59.1) |  |
| Parity, n (%) |  |  |  |  |
| 0 | 6 (6.1) | 6 (7.8) | 0 (0) | 0.281** |
| 1 | 19 (19.2) | 13 (16.9) | 6 (27.3) |  |
| 2 | 74 (74.7) | 58 (75.3) | 16 (72.7) |  |
| Performance status by ECOG score‡ | 1 (0-2) | 1 (0-2) | 1 (0-3) | 0.285 |
| Performance status by ECOG |  |  |  |  |
| 0 | 31 (31.3) | 24 (31.2) | 7 (31.8) | 0.188** |
| 1 | 35 (35.4) | 30 (39) | 5 (22.7) |  |
| 2 | 17 (17.2) | 14 (18.2) | 3 (13.6) |  |
| 3 | 14 (14.1) | 8 (10.4) | 6 (27.3) |  |
| 4 | 2 (2.0) | 1 (1.3) | 1 (4.5) |  |
| FIGO stage, n (%) |  |  |  |  |
| IIB | 63 (63.6) | 50 (64.9) | 13 (59.1) | 0.766** |
| IIIA | 2 (2.0) | 2 (2.6) | 0 (0) |  |
| IIIB | 34 (34.3) | 25 (32.5) | 9 (40.9) |  |
| Leukocytes, cells/μL‡ | 7870 (5540-9540) | 7960 (5620-9540) | 7080 (5020-9110) | 0.383¶ |
| Neutrophiles, cells/μL‡ | 4810 (3560-6800) | 4960 (3670-6800) | 4060 (3450-6450) | 0.421¶ |
| Lymphocytes, cells/μL‡ | 1510 (840-2210) | 1410 (800-2250) | 1605 (1040-1860) | 0.872¶ |
| Monocytes, cells/μL‡ | 470 (400-600) | 460 (400-640) | 500 (400-590) | 0.892¶ |
| Platelets, 10<sup>3</sup> cells/μL‡ | 306 (244-382) | 311 (244-367) | 296 (254-397) | 0.823¶ |
| RDW-CV, %‡ | 14.2 (13.3-16) | 14.1 (13.2-15.6) | 14.8 (13.8-17) | 0.186¶ |
| RDW-SD, fL† | 45.4 ± 6.1 | 44.7 ± 5.9 | 47.5 ± 6.4 | 0.074§ |
| Neutrophil to lymphocyte ratio‡ | 3.46 (2.23-6.11) | 3.5 (2.2-6.1) | 3.4 (2.2-4.5) | 0.780¶ |
| Lymphocyte to monocyte ratio‡ | 44.10 (41.60-49.80) | 3 (1.7-4.3) | 2.8 (2.1-3.6) | 0.772¶ |
| Platelet to lymphocyte ratio‡ | 14.20 (13.30-160) | 186.6 (148.4-339.3) | 221.8 (176-330.9) | 0.444¶ |
| Albumin, g/dL†€ | 3.74 ± 0.50 | 3.7 ± 0.5 | 3.8 ± 0.4 | 0.808§ |
| LMP1, n (%) |  |  |  |  |
| Negative | 87 (87.9) | 77 (100) | 10 (45.5) | <0.001** |
| Positive | 12 (12.1) | 0 (0) | 12 (54.5) |  |
| EBNA1, n (%) |  |  |  |  |
| Negative | 78 (78.8) | 77 (100) | 1 (4.5) | <0.001** |
| Positive | 21 (21.2) | 0 (0) | 21 (95.5) |  |
| Received treatment for cancer |  |  |  |  |
| Only RT | 25 (25.3) | 17 (22.1) | 8 (36.4) | 0.308** |
| QT/RT | 70 (70.7) | 57 (74.0) | 13 (59.1) |  |
| None | 4 (4.0) | 3 (3.9) | 1 (4.6) |  |
| Dead, n (%) |  |  |  |  |
| Alive | 41 (41.4) | 32 (41.0) | 9 (42.9) | 0.957 |
| Dead | 58 (58.6) | 45 (59.0) | 13 (57.1) |  |

n: number of patients; %: percentage of column; RDW: red blood cell distribution width; CV: coefficient of variation; SD: standard deviation; ECOG: Eastern Cooperative Oncology Group; FIGO: Fédération Internationale de Gynécologie et d'Obstétrique.
\* Unless otherwise indicated, p- values are for the comparisons of patients who were alive at the end of the study with those who died during follow-up using chi-squared test.
† Plus-minus values are mean ± SD.
‡ Values are median (percentile 25th-percentile 75th).
§ Two-sample t test with Satterthwaite's approximation formula to correct unequal variances.
¶ Mann-Whitney two sample test.
\*\* Fisher's exact test.
€ Only in 91 participants with complete data for albumin. Eight (8.1%) participants had missing data in albumin.

### Study endpoints

The main endpoint of this study was overall survival (OS), which was defined as the length of time (in months) from CC diagnosis until death by any cause. Those who still alive (without the event) until the end of the study period (July 2019) were treated as right-censored cases.

### Study variables

The marker of interest was the presence of EBV infection (EBV-positive), which was defined as a positive result for Epstein-Barr nuclear antigen 1 (EBNA-1) and/or Latent membrane protein 1 (LMP-1). Accordingly, the absence of EBV infection (EBV-negative) was defined as negative results for both LMP-1, and EBNA-1. We collected, from clinical records, data about the following sociodemographic and clinical covariates: age, parity, complete blood cell count (absolute numbers of leukocytes, neutrophils, lymphocytes, monocytes, platelets, red blood cell distribution width coefficient of variations [RDW-CV] and standard deviation [RDW-SD]), serum albumin, performance status, stage of cervical cancer and treatment received for cancer. Performance status was measured using the Eastern Cooperative Oncology Group (ECOG) scale. For cervical cancer staging, we used the International Federation of Gynecology and Obstetrics (FIGO) staging system (20). With the exception of the data regarding the management that the patients received to treat their cancer, all other covariates correspond to measurements that were registered in the patient’s medical records before or during the diagnosis of cervical cancer. The data was collected in a standardized case report form by an oncology resident with previous training in data collection.

### EBV analysis

#### Specimen collection

For EBV analysis, we collected tissue specimen obtained through incisional biopsy of the primary tumor site (cervix), which had been routinely fixed in formalin and embedded in paraffin blocks according to standard operating procedures at the Pathology Department of the Hospital Nacional Edgardo Rebagliati Martins. These samples were stored in the archives of the hospital until they were processed in 2019, average of five to six years after they were obtained. Only biospecimens with squamous cell carcinoma histology and whose films showed viable tumors were considered for the assay methods to detect EBV infection.

#### DNA isolation

Genomic DNA was purified from paraffin-embedded tissue sections using the QIAamp DNA FFPE Tissue Kit (QIAGEN, Valencia, CA) following the manufacturer instructions. Briefly, deparaffination was performed using two washes in xylene followed by two washes in ethanol.

Tissues were digested overnight at 65°C in 100 μl of lysis buffer containing 20 mg/ml of proteinase K. Genomic DNA was bound to QIAamp DNA columns and washed two times with QIAGEN washing buffers. The DNA was eluted in sterile water and the amount and quality of DNA were evaluated in an Eppendorf Biophotometer.

#### Polymerase chain reaction (PCR) assays

The PCR amplification was done in a total volume of 20 μl, containing 100 ng of DNA template. Thermal cycling was initiated at 95°C for 5 min, followed by 40 cycles of denaturation at 95°C for 30 secs, annealing at 61°C for 30 secs, extension at 72°C for 30 seconds, and a final extension at 72°C for 10 min. PCR products were detected using an EcoPCR Real-Time PCR instrument (Illumina, CA).

The presence of EBV genomic DNA was determined using the conditions and primer sequences for EBNA-1 and LMP-1 described by Ryan et al (21). The primers *EBNA-1* were as follows: EBNA1 Forward 5’-TACAGGACCTGGAAATGGCC-3’, Reverse 5’ TCTTTGAGGT CCACT GC CG-3’; LMP1 Forward 5’-CAGTCAGGCAAGCCTATGA-3’,Reverse 5’-CTGGTT CCGGTGG AGATGA-3’. Additionally, the amplification of human beta-actin gene (forward primer: 5’-ATCATGTTTGAGACCTTCAACAC-3’ and reverse primer: 5’-CATCTCTTGCTCGAAGTCCAG-3’) were used as an internal marker for the presence of intact genomic DNA.

The above PCR technique was chosen in view of its wide use in different EBV-related studies, its high reproducibility and based on a recent meta-analysis demonstrating a high sensitivity of this technique in the detection of EBV infection particularly when targeting EBNA-1 and LMP-1 (7).

### Statistical analysis

REMARK guidelines were followed in the analysis and reporting of this study (22). All statistical procedures were conducted using Stata/SE version 16.1 (StataCorp. 2019. Stata Statistical Software: Release 16. College Station, TX) for Windows 10 Pro x64 bits.

#### Exploratory data analysis and preliminary data presentation

We explored the distribution of numerical variables using analytical (mean, median, skewness, kurtosis, coefficient of variation) and graphical (histograms, box plot, quantile-quantile plot) analysis. We described numerical variables through means (standard deviations) or medians (percentile 25th and 75th), as appropriate. Categorical variables were described using absolute frequencies (n) and percentages (%). We estimated the median follow-up time using the observed period time and also the reverse Kaplan-Meier (K-M) method (23).

#### Association of EBV with established clinical and sociodemographic covariates

We explored associations of EBV with relevant sociodemographic and clinical variables. For continuous variables, we used a two-sample t-test with Satterthwaite’s approximation (for unequal variances) or the Wilcoxon rank-sum test, as appropriate. For categorical variables, we used the Chi-squared test or Fisher’s exact test, as appropriate.

#### Bivariable association between EBV with overall survival

Cumulative survival functions, time-specific survival rates (1-year and 5-year) were estimated using the K-M product-limit method. To gain a better comprehension of the univariable association between overall survival and EBV, we used different approaches. First, we modeled a simple Cox Proportional-Hazard (P-H) regression on a follow-up time axis to estimate the crude hazard ratio (cHR) of EBV to death (the complement of overall survival). Wald test was used for testing the bivariate association between EBV and mortality. After finding no graphical evidence of non-proportionality assumption through a visual inspection of the Schoenfeld residual against time plots, we compared the Kaplan-Meier cumulative survival curves using the log-rank test. Lastly, we estimated the risk difference (RD) of time-specific survival rates with 95% confidence interval p-values obtained by the pseudo-observations approach with the Stata “stpsurv” package (24).

#### Multivariable analysis

A multiple Cox P-H regression model was performed to examine the prognostic value of EBV after adjustment for the following well established prognostic factors and/or potentially confounding variables: age, parity (nullipara or monopara versus multipara), performance status by ECOG (0 versus 1 versus 2 versus 3), FIGO stage (IIB vs IIIA or IIIB), blood cell counts (neutrophils, lymphocytes, monocytes, platelets), RDW-SD, neutrophil-lymphocyte ratio (NLR), lymphocyte-monocyte ratio (LMR), and platelet-lymphocyte ratio (PLR). We adjusted only by variables that occur at or before the diagnosis of CC. The variables age, neutrophils, lymphocytes, monocytes, platelets, RDW-SD, NLR, LMR, and PLR were modeled as continuous predictors using a multivariable fractional polynomial modeling approach which avoids the drawbacks of categorization (25–27). We used the Stata package “mfp” for multivariable fractional polynomial modeling approach (MFP) (28). The non-linear transformations of continuous variables were modeled with four degrees of freedom using the closed test with a less stringent fractional polynomial selection type 1 error probability (α2 = 0.1). Categorical variables were modeled with one degree of freedom. The contribution to the model of each covariate was tested using a Wald test with a significance level of 0.05, but all covariates were adjusted into the model fixing the α1 of inclusion criteria in 1.00. The importance of EBV as a prognostic factor (our primary aim) was assessed via Wald test with a significance level of 0.05, the adjusted hazard ratio (aHR) for mortality (complement of survival), and its 95% confidence interval. All p values and 95% confidence intervals were estimated using a robust calculation of a variance-covariance matrix of standard errors clustered in each participant (29). Adjusted overall survival curves were estimated from the multivariable Cox P-H regression and 1-year and 5-years adjusted risk difference from a pseudo-observation approach using variables of the Cox model.

#### Checking model assumptions and internal model validation

We evaluated the linearity assumption during the model-building process through the MFP algorithm, which tests for nonlinearities and accounts for these using fractional polynomial transformations. Also, we used smoothed martingale residual plots to assess the accomplishment of the linearity assumption of the final multivariable model comparing it to a naïve multivariable model that had, by default, all continuous covariates in a linear transformation (see Supplementary Material). The proportional hazard assumption was assessed, firstly, using a multivariable fractional polynomial-time modeling approach (MFPT) that investigates for the presence of time-varying effects of all covariates included in the time-fixed multivariable model (30). We used the “stmfpt” package for MFPT, which allows us checking the presence of unacceptable departures of the PH assumption by comparing the fit improvements of the time-fixed model against models with interactions of a fractional polynomial transformation of time by covariate (30). Comparisons between time-varying alternative models were carried out using a closed test procedure with a significance level of 0.05 (30). After not finding statistical evidence of non-proportionality of hazards, we verified the PH assumption in the time-fixed multivariable model through the visual inspection of scaled Schoenfeld residual against time plots. The presence of outliers was assessed using deviance residuals against predictors plots, while influential points were investigated through DFBETAs against time plots. Lastly, we performed an interval validation of the multivariable model for assessing its stability creating 1000 bootstrap samples, and carrying out MFP Cox Proportional-Hazard regression in each resample. We used the Stata package “mfpboot” (31) and calculated the bootstrapped inclusion frequency (BIF) of EBV in resamples of the final multivariable model, as well as, resamples of an alternative multivariable model with different functional forms (fractional polynomials) of the continuous covariates.

## RESULTS

### Patient characteristics and follow-up

One-hundred eighty-nine patients were assessed for eligibility. A total of 99 (52.4%) patients with the diagnosis of squamous cell carcinoma of the cervix were included for analysis. Figure 1 summarizes the inclusion flowchart and Supplementary Table 1 details the reasons for exclusion. The median follow-up time in all participants, as calculated by the observed study period, was 2.57 years (range: 0.13 to 6.32 years), while the median follow-up time for the participants that remained alive was 4.69 years (range: 0.33-6.32 years). Additionally, the median follow-up time, as calculated by the reverse K-M method, was 5.22 years (percentiles 25^th^ to 75^th^: 3.67 to 5.66 years).

**Figure 1.**
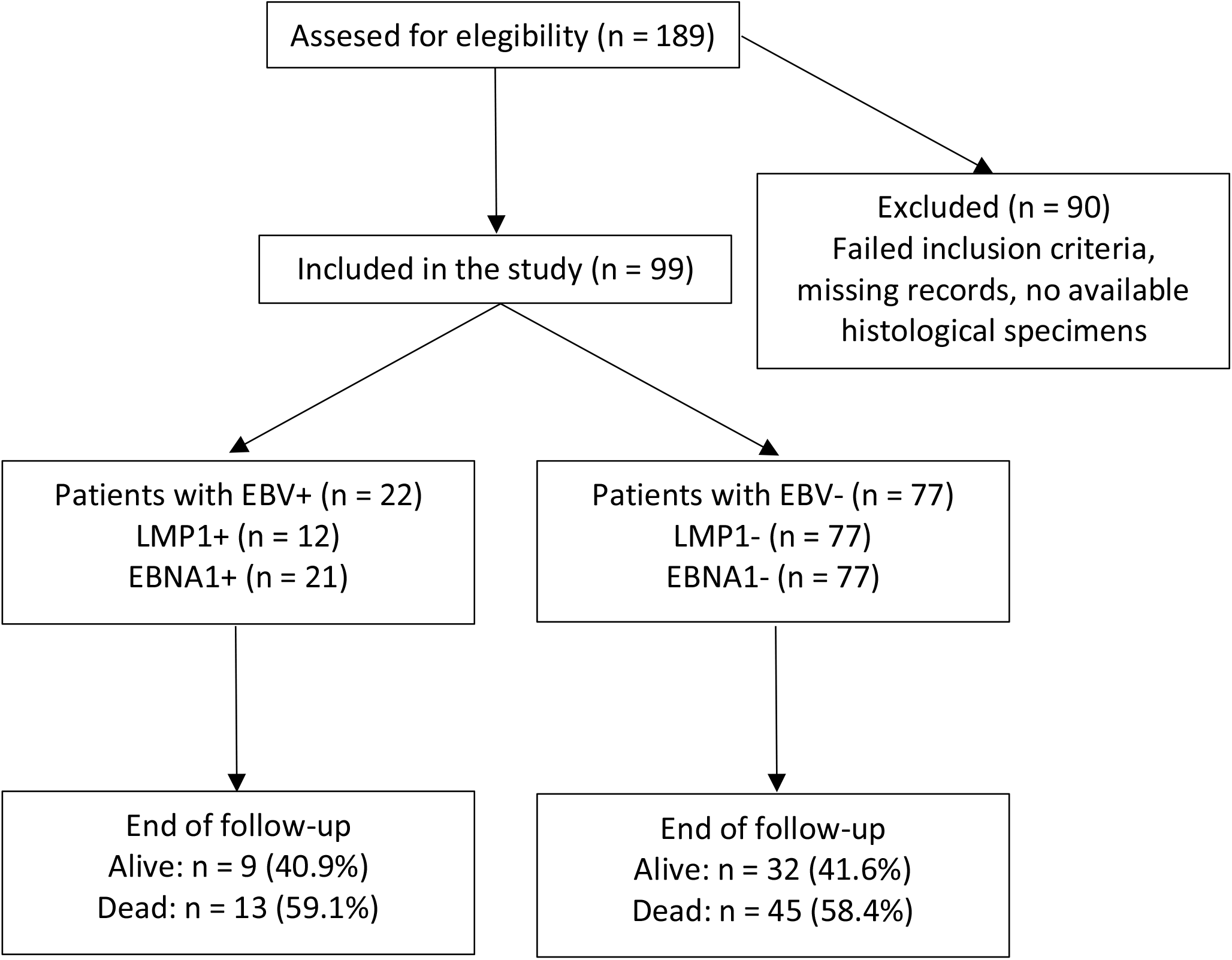
Flow-chart of participation in the study.

The patient characteristics are summarized in Table 1. The mean (SD) age was 55.1 years old, with a predominance of patients older than 45-years-old (66.7%, n = 67). Most patients had ECOG 0-1 (66.7%) and clinical stage IIB (63.6%) at the time of diagnosis. The prevalence of EBV in CC was 22.2% (n = 22) with EBNA-1 and LMP-1 positive results in 21.21% (n = 21) and 12.1% (n = 12), respectively. In the entire cohort, the 1-year overall survival was 79.5% (95% CI 70.0% to 86.2%) and the 5-year overall survival was 39.9% (95% CI 29.4% to 50.2%). At the conclusion of the follow-up period, more than half of the patients (58.6%, n = 58) had died.

### Bivariable association of EBV with established clinical and sociodemographic covariates

Results from the exploratory analyses showed similarities in the distribution of these continuous variables between positive and negative EBV patients (see Supplementary Figure 1). Concordantly, Table 1 showed no evidence of differences in the mean for age, RDW-SD and serum albumin level according to EBV status, and also no evidence of differences in the median for ECOG score, leucocytes, neutrophils, lymphocytes, monocytes, platelets, RDW-CV, NLR, LMR, and PLR according to EBV status. Similarly, there was no evidence of associations between EBV and the remaining covariates such as parity, and FIGO stage (Table 1).

### Association of EBV with overall survival

Figure 2 shows the crude and adjusted survival curves, respectively. There were differences in adjusted survival rates by EBV status (Figure 2B) although without evidence of differences in the crude analysis (Figure 2A). Thus, while the crude comparisons of 5-years OS reveal non-statistically significant RD of −8.9% (95% CI −35.8% to 18.0%; p = 0.518), there were a large difference in the adjusted 5-year OS rates (RD = −29.5; 95%CI −53.4 to −5.6; p = 0.016) (Table 2).

**Table 2.** Relationship between EBV and OS.

|  | No. of Patients | No. of Deaths | Overall Survival |  |
| --- | --- | --- | --- | --- |
|  |  |  | 1-year (95% CI) | 5-year (95% CI) |
| **EBV-positive** | 22 | 13 | 81.8 (58.5 to 92.8) | 45.0 (23.9 to 64.1) |
| **EBV-negative** | 77 | 45 | 78.8 (67.7 to 86.4) | 37.8 (25.7 to 49.8) |
| **Crude RD, %** |  |  | -3.1 (-22.0 to 15.8); p = 0.746 | -8.9 (-35.8 to 18.0); p = 0.518 |
| **Adjusted RD, %** |  |  | -5.6 (-23.4 to 0.12); p = 0.538 | -29.5 (-53.4 to -5.6); p = 0.016 |

OS: overall survival; RD: Risk Difference

**Figure 2.**
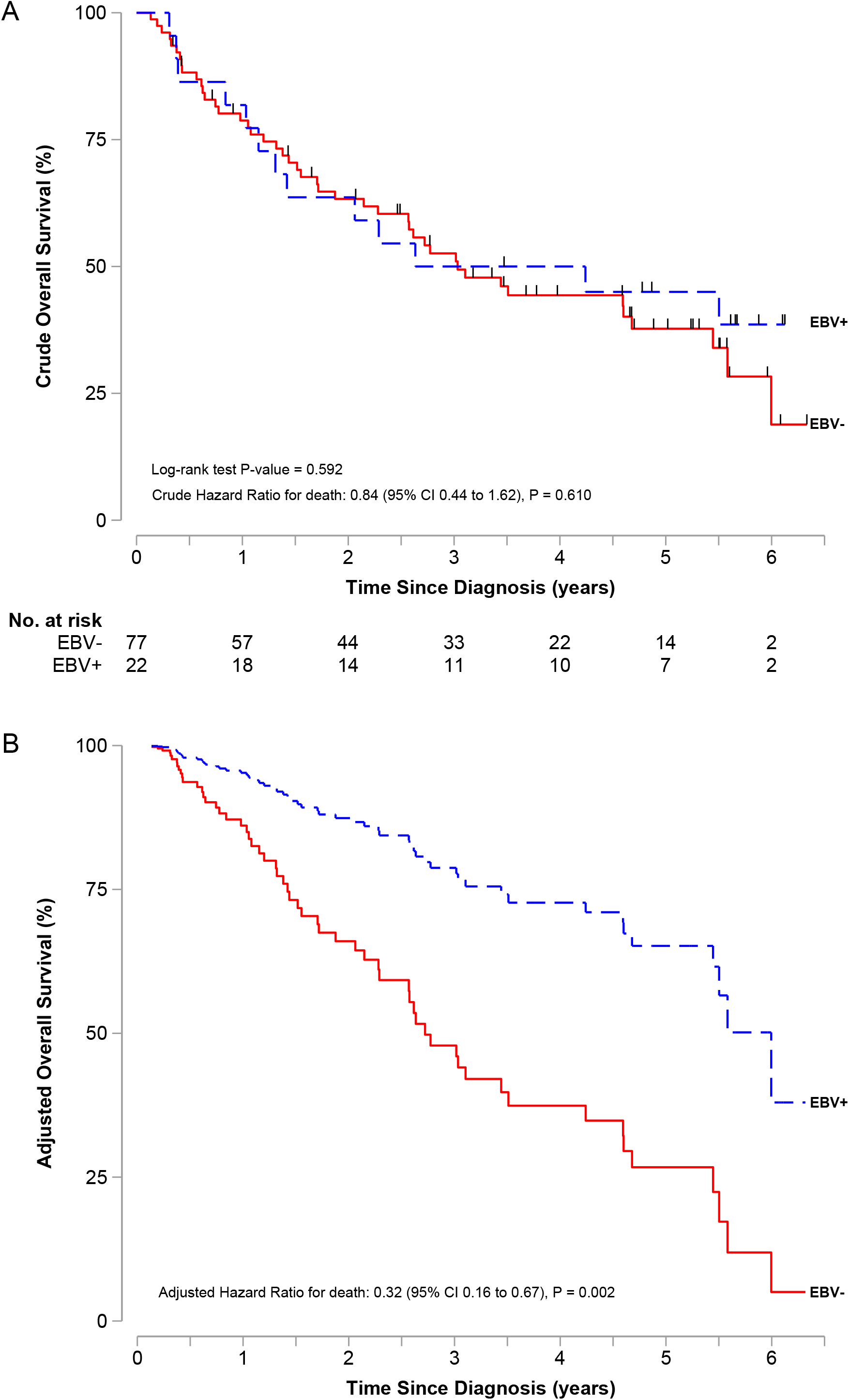
(A) Crude and (B) adjusted overall survival in patients with EBV positive and EBV negative. Hash marks show censoring time.

Concordantly, the bivariable Cox P-H regression, reported in Table 3, showed that EBV-positive patients had 16% less instantaneous risk of death compared to the EBV-negative patients, although this association was not statistically significant (cHR = 0.84; 95% CI 0.44 to 1.62; p = 0.610). However, after adjusting for established prognostic factors and potential confounders factors (see Table 3), EBV-positive patients had 68% less instantaneous risk of death compared to EBV-negative patients (aHR = 0.32; 95% CI 0.16 to 0.67; p = 0.002).

**Table 3.**
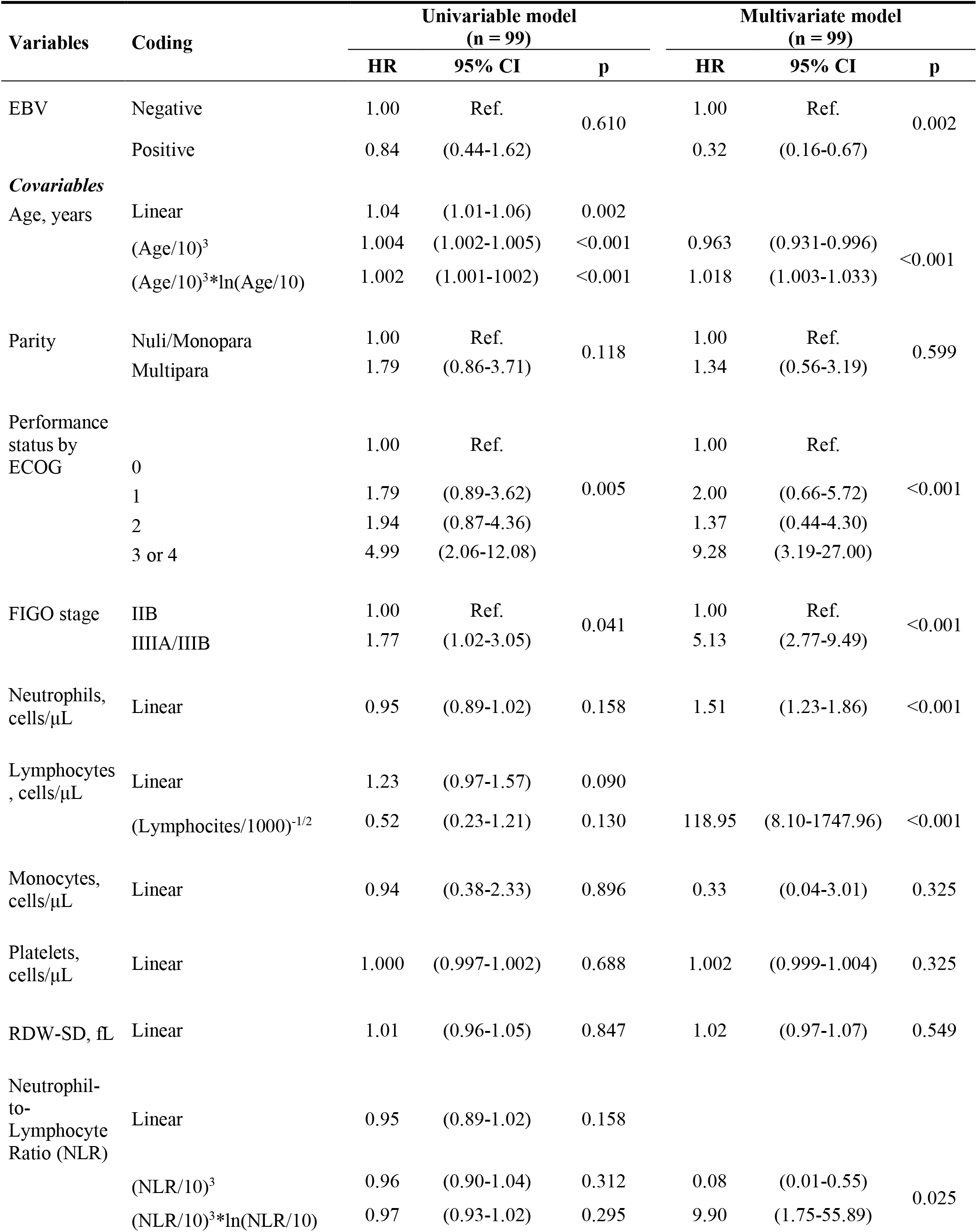
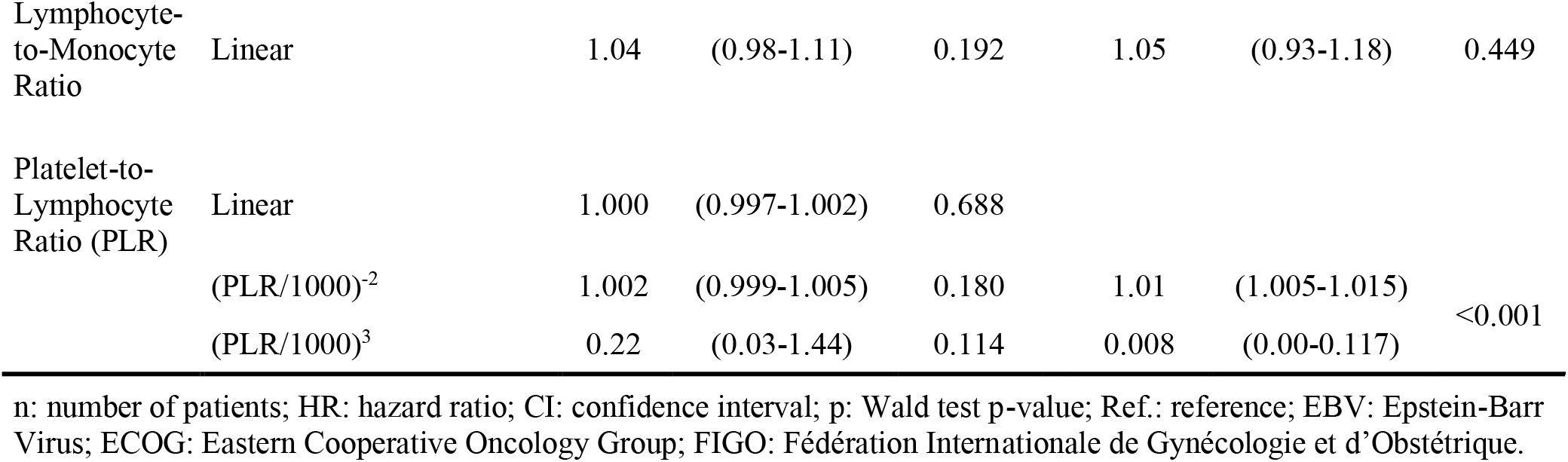
Association between clinical-pathological features and mortality.

### Checking model and stability analysis results

There was no evidence of time-varying effects in any covariate included in the fixed-time model, as evaluated with the MFPT approach. Accordingly, the scaled Schoenfeld residual against the time plot did not show evidence of departures of the proportionality of hazards assumption (see Supplementary Figure 2). Smoothed martingale residuals plots did not show evidence of violations of the linearity assumption in our final multivariable model (Supplementary Figure 3). No influential points for adjusted HR estimates on EBV status were found, but some potentially influential points for NLR, PLR, lymphocytes, and monocytes were observed (Supplementary Figure 4). A more detailed investigation using alternative models that excluded observations with these potentially influential points showed consistent estimates of adjusted HR for EBV (data not shown).

The stability analysis for internal validation showed a relatively high BIF of 70.8% for our marker of interest (EBV) demonstrating that it was a strong predictor of overall survival. Consistently, all bootstrapped aHR for EBV were statistically significantly less than the null value (aHR = 1.00). However, we found relatively moderate BIF in the fractional polynomial degree 2 (FP2) transformations for PLR (BIF = 55.4%), and age (BIF = 55.0%). Other continuous covariates showed BIFs of 100%, except for the FP2 transformation of NLR that showed a relatively low BIF of 38.7%. Furthermore, all covariates showed the inconsistent direction of the aHR (with values > or < of 1.00). A sensitivity analysis performed choosing a more stable fractional polynomial transformation for PLR showed that the aHR for EBV was the same (aHR = 0.32; 95% CI 0.15 to 0.70; p = 0.005) and its BIF was slightly increased to 72.0%. However, three of 1000 bootstrapped resamples had inconsistent aHRs characterized by values higher than 1.00. All these findings revealed the presence of instability in the non-linear transformation for these variables, apparently with a little impact on the stability of the aHR for the marker of interest, EBV.

## DISCUSSION

In this study, we hypothesized that the presence of EBV infection in cervical tissue could have produced a negative prognostic effect in CC, this based on previous findings reported in patients with aggressive B-cell lymphomas where EBV infection was correlated with worse survival rates (10,18), and also based on previous reports showing that EBV can enhance and accelerate HPV integration to promote tumor progression (32). However, in our cohort of

Peruvian women with cervical cancer, EBV infection was associated with an improved OS compared to EBV-negative individuals. To the best of our knowledge, this is the first study assessing the relationship of EBV infection and its impact on overall survival in patients with CC.

Although our findings seem counterintuitive, knowledge about the role of EBV as a prognostic factor for better OS in some cancers is not new, since it has also been described in EBV-associated gastric carcinoma (EBVaGC) (11,33) and undifferentiated/non-keratinizing nasopharyngeal carcinoma (NPC) (12,34). EBVaGC and undifferentiated/non-keratinizing NPC are unique entities associated with an overall good prognosis. The largest study on EBVaGC reported lower mortality rates (HR, 0.72; 95% CI 0.61 to 0.86) and better OS (8.5 years versus 5.3 years in EBV-positive versus EBV-negative patients, respectively, p < 0.001) (11). In NPC cases, a better prognosis was found in EBV-positive non-keratinizing carcinomas compared to EBV-negative carcinomas (36 months versus 16 months, respectively, p = 0.004) (12). Both EBV-positive neoplasms possess a tumor microenvironment composed of a dense immune infiltrate (33,35–38). This increased peritumoral and intratumoral immune cells infiltrate is composed by cytotoxic CD8+ T lymphocytes and dendritic cells that would trigger an immune response that strengthens the host immune system and will enhance anti-tumor activities (35,38,39).

Based on our findings, we believe EBV-associated CC could have a similar behavior of what is seen in EBVaGC and undifferentiated/non-keratinizing NPC. However, the precise mechanism of how EBV infection results in better survival in patients with CC remains to be elucidated. A study demonstrated that there is not only an enrichment of a T-cell immune signature, but also an increased B-cell signature in HPV-positive and EBV-positive tumors separately, meaning that both T-cell and B-cell tumor-infiltrating lymphocytes (TILS) present in a tumor lead to a better survival (37). Moreover, both viruses are associated with a high expression of immunosuppressive genes such as PD-1 (programmed cell death protein 1), PD-L1 (programmed death-ligand 1), PD-L2 (programmed death-ligand 2), CTLA-4 (cytotoxic T-lymphocyte antigen 4), Tim-3 (t-cell immunoglobulin mucin-3), LAG3 (lymphocyte Activation Gene-3), and ICOS (inducible co-stimulator)(37). Furthermore, previous studies have shown that the presence of immune-inflammatory infiltrate enhance the response to checkpoint inhibition (anti-PD-1/PD-L1 and anti-CTLA) and lead to better survival in melanoma (40), meaning that the inflammatory infiltrate has a positive predictive role to the release of immune checkpoints by the inhibitors, increasing in this way the lymphocyte proliferation and the antitumor effect (41).

An additional finding in our study is that nearly 1 of 5 women (22.2%) was positive for EBV in our cohort of CC. This prevalence is less of what has been reported in a previous meta-analysis made from 25 studies which revealed an overall pooled prevalence of EBV of 33.4% (7). Furthermore, our study did not find any differences related to epidemiological and clinical features between EBV-positive and EBV-negative individuals which is inline of what has been described in previous report (14).

During recent years, HPV/EBV co-infection has been an interesting subject of discussion in regards to the impact of this interaction on overall survival. For instance, in NPC it has been shown that the presence of both EBV and HPV infection were independent positive prognostic factors for survival (34). Also, a recent study from Finland showed a better prognosis in HPV-positive/EBV-positive co-infection than in patients with HPV-negative/EBV-positive or HPV-negative/EBV-negative oropharyngeal cancer (42). However, in the case of the co-infection between H. pylori and EBV in gastric cancer, it is known that this interaction increases the oncogenic potential and development of gastric cancer, however, it is unclear how this interaction influences in a patient’s outcomes and survival (43). Regarding CC, the relationship that occurs between both viruses has been less studied and therefore is one of the least understood among virus-related cancers to date.

Contrary to our hypothesis, it has been proposed that EBV can accelerate the integration of the HPV genome in cervical cells (13), which could then lead to increased instability, facilitate oncogenic effects and could promote progression and metastasis (17,32). This proposed mechanism consists that viral EBV oncoproteins (EBNA1, LMP1, LMP2A) and high-risk HPV oncoproteins (E5, E6, and E7) may be present and cooperate with the initiation and/or amplification of epithelial-mesenchymal transition (EMT) which is a hallmark of progression and metastasis (17,32). Following this, since it is known that EBV and HPV oncoproteins share several signaling pathways, such as JAK/STAT/SRC, β-catenin, RAS/ MEK/ERK and/or PI3k/Akt/mTOR, it has been postulated that a cooperative activation could be the main mechanism for metastatic progression through EMT amplification (17,32,44,45). However, the exact mechanism of interaction between both viruses and its role in CC carcinogenesis is not completely understood and more studies are needed. Despite the reasonable biological plausibility of the described theoretical proposals, they do not seem to agree with the empirical evidence in the case of CC. According to these, we would expect to see worse mortality associated with EBV-positive in patients with CC, whereas our findings find just the opposite. This does not mean that such mechanisms do not exist, but rather that their role may be less than that of other antitumor activity mechanisms, such as TILS. Future studies could assess how these mechanisms interact and counter-regulate in the complex cervical cancer microenvironment, promoting, and/or blocking tumor development.

The fact that in our study the association between EBV infection and overall survival was such strong and significant in the multivariable analysis (yet not statistically significant in the crude analysis) had two main explanations, and one direct practical implication. Firstly, these findings suggest the existence of important confounding factors in the hypothetical causal relationship between EBV and overall survival in CC patients. Thus, only after controlling for confounding factors we were able to have a better estimate of the true relationship between both variables. Secondly, an insufficient power for detecting significant associations could partially explain our results, considering that the crude association revealed a 16% less instantaneous risk of death for EBV-positive comparing with EBV-negative and that we had a limited sample size of only 99 patients. By last, one clinical practical implication of this apparent contradiction between crude non-significant versus adjusted significant results is that although testing for EBV infection in clinical practice would not be an appropriate prognostic maker to be used alone, perhaps, it could be a promissory candidate for improving existent prognostic models commonly used such as the FIGO system (20), and new models that are being incorporated such as the neutrophil-to-lymphocyte ratio and platelet-to-lymphocyte ratio (46,47). Further studies, with well-powered sample sizes and a better consideration of confounding variables should be done to specifically assess this possible causal relationship or simply for a better determination of the prognostic value of EBV status on overall survival in CC patients.

As any retrospective study, our study has some limitations that deserve to be discussed. Firstly, the retrospective design of this study limited the availability of complete data. Thus, during the adjusted analysis we decided not to use serum albumin level since a relatively important proportion of this data was missing (8.8%). Aside from this variable, all the reviewed registries had complete data to perform an appropriate analysis of the remaining covariates. Secondly, the risk for selection bias cannot be excluded from this study, especially since we had to exclude 61 otherwise eligible patients due to insufficient tissue sample needed for PCR analysis. However, exclusion of CC patients most likely happened in a random, non-selective fashion because cervical tissue losses were primarily due to deterioration of the paraffin-embedded blocks over time. Therefore, we expect a lesser impact in the selection of our patients and in our estimates of aHR for EBV. Thirdly, our study was conducted in a single center that only attend insured patients through the Social Security System in Peru, thus generalizability cannot be guaranteed. However, this hospital is a highly specialized cancer referral center concentrating most cases of cervical cancer in Peru, therefore, these findings could be reasonably extrapolated to a wider sector of this insured people. Lastly, our results have not been confirmed in an external cohort of patients that meet similar criteria; however, we did perform an internal validation based on bootstrapping, which demonstrated a relatively high inclusion frequency of EBV infection and values of aHR consistent with a protective effect. Therefore, our promissory results are preliminary and must be replicated in a derived prospective cohort to validate them.

A main strength of our study is the efficient use of data through continuous modeling, which increases the statistical power and avoids the arbitrary election of cut-off for the biomarkers and the resultant drawbacks of categorization. Also, another strength was that we used the PCR technique which is a highly sensitive method to detect EBV in tissues of seropositive individuals (21). Although the gold standard for detection of EBV (EBER study) consists in detecting the latent EBV infection, this technique is unable to detect cases when the virus is in lytic phase and in consequence this method has a lower sensitivity with more probability to have false negative results (48).

In light of these findings, one possible implication of demonstrating the role EBV infection and its impact on survival is that we could apply a de-escalation therapy approach according to EBV status. For instance, EBV-positive patients could be treated with a radiotherapy-only approach preventing unnecessary toxicity from chemotherapy. Likewise, EBV-negative patients could benefit more from a concurrent approach such as chemoradiotherapy. Another possible implication is the development of clinical trials evaluating the role of immune checkpoint inhibitors in patients with locally advanced or metastatic EBV-positive CC. This based on data demonstrating a presumed positive effect of immune checkpoint inhibitors on viral-associated cancers, especially in the squamous cell carcinoma subtypes (49–51)

In conclusion, our study shows, for the first time, that the detection of EBV infection in cervical cancer tumor specimens was associated with improved overall survival compared to EBV-negative individuals. The evaluation of EBV status could be used as a clinical prognostic biomarker and to improve currently available prognostic models. Future prospective studies will be needed to validate these theories.

## Data Availability

The data supporting the findings of this study are available in the Research Unit of the Oncology Department of the Hospital Nacional Edgardo Rebagliati Martins computer files, but restrictions apply to the availability of these data, which were used under the current study license and are therefore not publicly available. However, the data are available to the authors upon reasonable request and with the permission of the Research Unit.

## 2. Author Contributions

DC and BB contributed the conception, design, and supervision of the study. AC collected the information on the clinical records and organized the data. JV collected the patient samples and confirmed the pathology. MLI performed the RT-PCR for EBV in cervical tissues. PSB performed the quality control checks of the database and the statistical analysis. DC, BB, PSB, LM, JV, MLI, AY, AC, and AF wrote the manuscript. All authors contributed to manuscript revision, read, and approved the submitted version.

## 4. Acknowledgments

We thank the CDX Molecular company providing the RT-PCR platform. The center is located in 309 Javier Prado Avenue, San Isidro district, Lima-Peru.

## 5. Supplementary Material

The Supplementary Material for this article can be found online at:

